# Self-collected oral, nasal and saliva samples yield sensitivity comparable to professional-collected oro-nasopharyngeal swabs in SARS-CoV-2 diagnosis

**DOI:** 10.1101/2021.04.13.21255345

**Authors:** Maximilian Gertler, Eva Krause, Welmoed van Loon, Niklas Krug, Franka Kausch, Chiara Rohardt, Heike Rössig, Janine Michel, Andreas Nitsche, Marcus A. Mall, Olga Nikolai, Franziska Hommes, Susen Burock, Andreas K. Lindner, Frank P. Mockenhaupt, Ulrich Pison, Joachim Seybold

## Abstract

**Introduction:** Containment of the COVID-19 pandemic requires broad-scale testing. Laboratory capacities for real-time-PCR were increased, and are complemented by Ag-tests. However, sample-collection still requires qualified personnel and protective equipement, may produce transmission to others during conduct and travel, and is perceived uncomfortable. We tested sensitivity of three simplified self-sampling techniques compared to professional-collected combined oro-nasopharyngeal samples (cOP/NP).

**Methods:** From 62 symptomatic COVID-19 outpatients, we obtained simultaneously three self- and one professional-collected sample after initial confirmation in a testing centre: (i) combination swab (tongue, cheek, both nasal vestibula, MS, (ii) saliva sponge combined with both nasal vestibula, SN, and (iii) gargled tap water, GW, (iv) professionally-collected cOP/NP (standard). We compared the results of SARS-CoV-2 PCR-assays detecting E-gene and ORF1ab for the different sample types and performed bivariate statistical analysis to determine the variables reducing sensitivity of the self-collecting procedures.

**Results:** SARS-CoV-2 RNA was detected in all 62 professionally-collected cOP/NP. MS and SN samples showed a sensitivity of 95.2% (95%CI 86.5-99.0) and GW samples of 88.7% (78.1-95.3). Compared to the median *ct*-values of cOP/NP samples for *E-gene* (20.7) and *ORF1ab* (20.2) these were higher for MS (22.6 and 21.8), SN (23.3 and 22.3), and for GW (30.3 and 29.8).

For MS and SN samples but not for GW specimens, false negativity in bivariate analysis was associated with non-German mother-tongue, number of sampling errors, and with symptom duration. For symptom duration of ≤8 days, test sensitivity for SN samples was 98.2% (95%CI 90.4-100.0) and for MS 96.4% (95%CI 87.7-99.6) and drops after day 8 below 90%.

**Discussion:** The study is limited to sensitivity of self-collection in symptomatic patients. Still, in this group, self-collected oral/nasal/saliva samples are reliable alternatives to professional-collected cOP/NP samples, if symptom duration does not exceed eight days and operational errors are minimized. Self-sampling could contribute to up-scaling of safe and efficient testing.

## Introduction

Containment of the current COVID-19 pandemic(Lu et al., 2020, Zhu et al., 2020) requires broad-scale testing capacities(Zhu and Wong, 2020) for patients, potentially contagious persons and groups at risk of infection. Laboratory capacities for real-time reverse transcriptase polymerase chain reaction (rtRT-PCR) have been significantly increased in many countries, and are complemented by novel rapid test devices based on antigen detection(Rai et al., 2021). Still, professional-collected (oro-)nasopharyngeal samples are considered the gold standard(Marty et al., 2020, Pan et al., 2020). However, this approach remains challenging considering the needs of qualified medical personnel and protective equipment as well as the risk of potential virus transmission to health care workers or others at testing-sites(Zhu and Wong, 2020). Moroever, (oro-)nasopharyngeal sampling is being perceived as uncomfortable, and possibly deterring, by many patients. Simplified sampling techniques may help overcoming these limitations. Reliable self-collecting procedures could be home-based and thus, contribute to reduced virus transmission due to more rapid diagnosis and reduced mobility of potentially contagious persons. Self-collected samples from the oral cavitiy, e.g. saliva or from the nasal vestibule (anterior nasal cavitiy) are therefore being investigated as non-invasive, more comfortable and less resource-intensive alternatives and show variable reliability(Fernandez-Gonzalez et al., 2021, Tu Yuan-Po et al., 2020, Tu Y.P. et al., 2020).

We performed a prospective manufacturer-independent sensitivity study in symptomatic SARS-CoV-2 positive outpatients to examine whether combinations of simple self-collection technics may be reliable alternatives to professional-collected pharyngeal sampling. We obtained four simultaneous samples for rtRT-PCR testing from these patients: one professional-collected, oro-nasopharyngeal swab sample and three self-collected specimens using different simplified sampling procedures from more distal locations in the upper respiratory tract. Herein, we present the sensitivities using self-collected samples as compared to the gold standard.

## Methods

### Study design and Participant Enrolment

We calculated 60 patients with confirmed SARS-CoV-2 infection as necessary to compare the sensitivity of rtRT-PCR assays in self-collected versus professional-collected samples assuming a true sensitivity of 98%.

Between 7th December 2020 and 11th January 2021, we prospectively enrolled 62 SARS-CoV-2 infected outpatients into the present sudy. On the day before enrolment, all patients had attended the central testing site of Charité – Universitaetsmedizin Berlin (Maechler et al., 2020), with symptoms compatible with COVID-19. Professional-collected, combined oro-nasophayryngeal swabs were subjected to RT-PCR at the central Charité laboratory. Upon results communication and counselling *via* telephone, study participation was offered to patients.

Symptomatic patients were eligible in case of a confirmed RT-PCR test result not older than 24 hours before phone contact, and location of residence enabling a home visit on the same day.The study was reviewed by the ethics committee at Charité-Universitätsmedizin Berlin, Germany (EA2/192/20), and written informed consent was obtained prior to study entry.

A medical study team visited the participants and interviewed them on perception of the below procedures, their professional and linguistic background and competences, as well as on prior experience with swabs. Medical professionals collected an oro-nasopharyngeal swab as reference sample. Next, the team handed written instructions and necessary materials for three self-collecting procedures. To assess independent patient-self-collection, the collections were observed without any additional verbal instructions or intervention, and performance and irregularities were documented by the study team. The three procedures of self-collection included (i) MS (multi-swab): a combination swab from the tongue, the inner cheek and both nasal vestibules (insertion 2-3 cm, twisting 4x), (ii) SN (saliva-nasal): insalivating of the swab for 10-15 sec. before swabing both nasal vestibules (insertion 2-3 cm, twisting 4x), and (iii) GW (gargle water): collection of 10 ml of gargled tap water into a plastic container (Sarsted^R^, L494-9). Swabs used were nylon-flocked applicators with 1 ml of Amies preservation medium (ESwab® Copan, Italy).

### Laboratory analyses

For this study, we performed a quadruplex RT-PCR assay to simultaneously detect the *E-gene* and *ORF1ab* of SARS-CoV-2, the *c-myc* gene representing human nucleic acid, and *KoMa*, an artificial sequence that has no significant homology to any sequence in GenBank. Total RNA was extracted with the QIAamp Viral RNA Mini Kit (Qiagen, Hilden, Germany). PCR was performed using the AgPath-ID™ One-Step RT-PCR Reagents kit (Applied Biosystems, Foster City, CA USA) on a Bio-Rad CFX96 device. Cycling conditions were: 45°C for 15 min, 95°C for 10 min followed by 45 cycles of 95°C for 15 s and 60°C for 30 s. The *E-Gene* primer and probe sequences were taken from Corman et al.(Corman et al., 2020). The SARS-CoV-2 specific *ORF1ab* assay was designed based on the 72 sequences that were available at the time from Michel et al.(Michel et al., 2020). *KoMa* and human cell *c-myc* were detected to serve as quality and internal amplification (*KoMa*) control for potential inhibitors of RT-PCR and the respective sampling technique(Kirchner et al., 2010). The cycle threshold-value (ct-value), i.e. the PCR cycle at which the fluorescence signal crosses the detection threshold, was determined for each target sequence.

Probit analysis revealed the limit of detection for the quadruplex PCR under the above described conditions as 28.7 genome copies for the *E-Gene* assay and 32.0 genome copies for the *ORF1ab* assay. Tests with signals that crossed the detection threshold were considered positive. All samples were measured on the day of collection by using 140 µl aliquots for RNA extraction, respectively.

### Statistical analyses

Descriptive statistics used proportions, means ± standard deviation (SD), or medians with interquartile ranges (IQR), as applicable. Categorial variables were compared by two-tailed Fisher’s exact test, numeric variables by a Mann Whitney U-test, and paired numeric data by a Wilcoxon Signed Rank test. Sensitivity and 95% confidence interval (CI) were calculated for each sampling method.

We assessed the linear dependence between *ct*-values of the professional collected swabs and *ct*-values of the self-collecting sampling methods by target gene, using Pearson correlation (r). A binominal regression model was used to determine the variables reducing sensitivity of the self-collecting procedures (negative RT-PCR) compared to the correspondent professional collected sample. All computations were performed using “R” version 3.6.3 for all analyses. *P*<0.05 was considered to reflect statistical significance.

## Results

### Comparison of sensitivities and *ct*-values by sampling technique

All 62 participants provided three self-collected samples (*n* = 186) in addition to the 62 newly obtained professional-collected oro-nasopharyngeal specimens. Half of the patients were female, and their median age was 31.5 (range, 17-66). The medium time between onset of symptoms and enrolment was four days (range, 2-15). Symptoms and other clinical variables are presented in table 1.

**Table 1:** Patient characteristics and symptoms.

|  | All patients (N=62)<br>% (n) or median (range) |
| --- | --- |
| Female | 50% (31) |
| Age (years) | 31.5 (17.0, 66.0) |
| Shortness of breath | 6.5% (4) |
| Chest pain/Chest tightness | 0% (0) |
| Fever in the last 48 hours | 30.6% (19) |
| Chills | 38.7% (24) |
| Fatigue | 80.6% (50) |
| Body aches | 66.1% (41) |
| Cough | 62.9% (39) |
| Rhinorrhea | 67.7% (42) |
| Diarrhea | 21% (13) |
| Sore throat | 41.9% (26) |
| Headache | 75.8% (47) |
| Impaired smell and taste | 50% (31) |
| Time between test and symptom onset (days) | 4.0 (2.0, 15.0) |
| Chronic lung disease | 9.7% (6) |
| Diabetes I/II | 1.6% (1) |
| Cardiovascular disease | 8.1% (5) |
| Obesity | 6.4% (4) |
| Contact to a confirmed SARS-CoV-2 case | 33.9% (21) |
| Time between test and last contact (days) | 6.0 (1.0, 10.0) |
| German NOT as first language | 30.6% (19) |

All 62 professional-collected samples tested positive for both SARS-CoV-2 target genes, the *E-gene* and the *ORF1ab*. In all samples, regardless of sampling technique, *c-myc* was detected, indicating that all contained human cells. This was done using the *c-myc* PCR assay in a singleplex reaction, as samples which are positive in the *E-gene* and *ORF1ab* PCR assays do not always allow the amplification of the *KoMa* and *c-myc* controls in the quadruplex reaction. No signs of PCR inhibition were detected.

Detection sensitivities for *E-gene* and *ORF1ab* differed depending on the self-collecting procedure. For *E-gene*, MS samples were positive in 93.5% (range: 84.3-98.2), SN samples in 95.2% (86.5-99.0), and GW samples in 87.1% (76.1-94.3). For *ORF1ab*, both MS and SN samples were positive in 91.9% (82.2-97.3), and GW samples in, 88.7% (78.1-95.3). Defining a sample as SARS-CoV-2 positive if either *E-gene* or *ORF1ab* was detected, MS and the SN samples showed a sensitivity of 95.2% (86.5-99.0) and GW samples of 88.7% (78.1-95.3).

Figure 1 presents *ct*-values of all samples tested positive for *E-gene* and for *ORF1ab*. Median *ct*-values of the professional collected samples was 20.7 for *E-gene* and 20.2 for *ORF1ab*. Median *ct*-values of the self-collected samples were slightly but significantly (p<0.05) higher for MS (22.6 and 21.8) and SN (23.3 and 22.3), and substantially so for GW (30.3 and 29.8).

**Figure 1.**
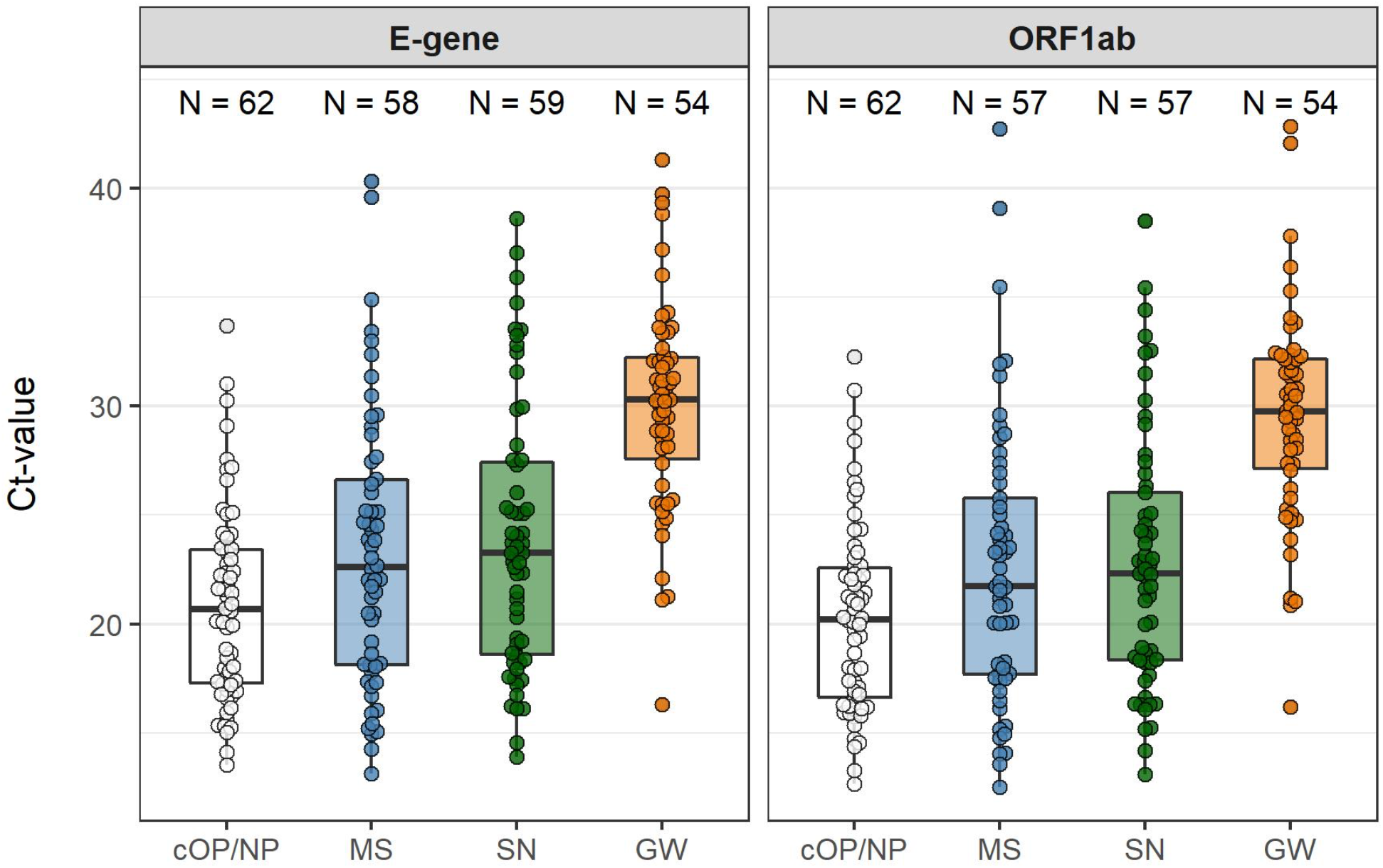
Ct values for *E-gene* and the *ORF1ab* of all positive samples by type of sample collection: professional-collected cOP/NP sample (white), patient-collected samples MS (blue), SN (green), and GW (orange) The boxes in the plot depict the 25th, 50th and 75th percentiles.

For both target genes, *ct*-values of all sampling techniques were significantly correlated with their respective *ct*-values for the cOP/NP (p<0.001 for all comparisons), with the strongest correlation for the *E-gene* in MS (r=0.77) and SN (r=0.73) (Figure 2).

**Figure 2.**
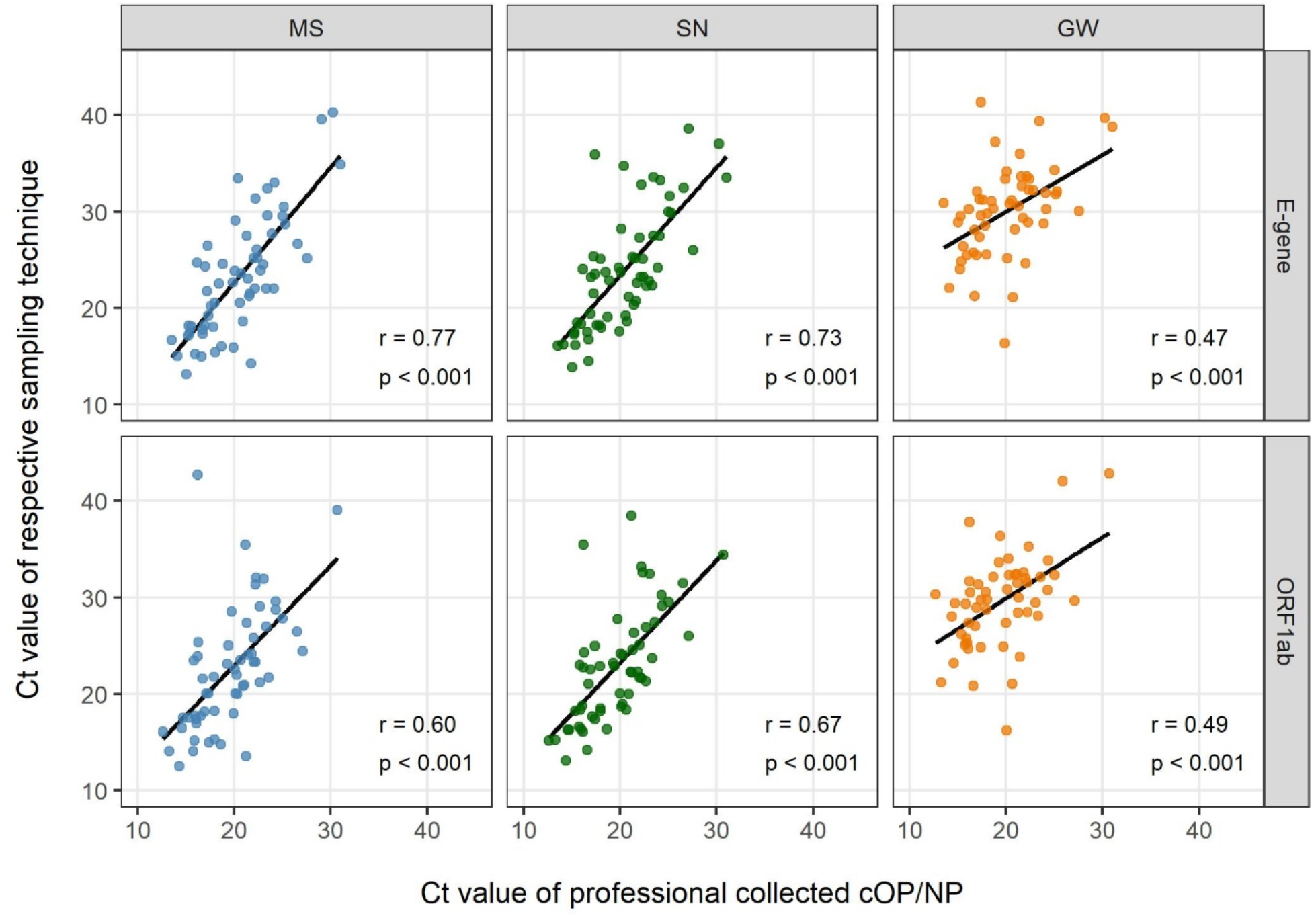
Ct-values of each patient-collected sample type (MS, SN, GW) compared with the ct-value of the diagnostic standard (medical professional-collected OP/NP sample, x-axis) shown for the used two target genes. MS (Multi-swab – blue), SN (Saliva-nasal – green), GW (gargle water – orange)

### Factors associated with false-negative results in patient-collected samples

In three (4.8%) MS and SN samples as well as in seven (11.3%) GW specimens, none of the two SARS-CoV-2 genes were detected. These false negative results were associated with high *ct*-values, i.e. low viral loads, in the corresponding professional-collected swab (Table 2). For MS and SN samples, but not for GW specimens, false negativity was also associated with a non-German mother-tongue, the number of sampling procedure mistakes, and as a trend, with symptom duration.

**Table 2:** Comparison of variables between negative and positive test result in patient collected samples separated by collection method.

| Sample type | Variable | Negative | Positive | p-value |
| --- | --- | --- | --- | --- |
| MS | N | 3 | 59 |  |
|  | Ct value of professionally collected cOP/NP, median (IQR) | 27.2 (27.1, 30.4) | 20.4 (17.2, 22.9) | 0.008 |
|  | Female | 1 (33.3%) | 30 (50.8%) | 1.000 |
|  | Age in years, median (range) | 48.0 (35.0, 53.0) | 31.0 (17.0, 66.0) | 0.064 |
|  | Number of symptoms, median (range) | 7.0 (7.0, 7.0) | 5.0 (2.0, 10.0) | 0.335 |
|  | Symptom interval in days, median (range) | 11.0 (4.0, 15.0) | 4.0 (2.0, 12.0) | 0.083 |
|  | German is not first language | 3 (100.0%) | 16 (27.1%) | 0.026 |
|  | No. of mistakes during sampling procedure, median (range) | 2.0 (2.0, 3.0) | 0.0 (0.0, 3.0) | 0.005 |
| SN | N | 3 | 59 |  |
|  | Ct value of professionally collected cOP/NP, median (IQR) | 29.1 (28.1, 31.4) | 20.4 (17.2, 22.9) | 0.006 |
|  | Female | 1 (33.3%) | 30 (50.8%) | 1.000 |
|  | Age in years, median (range) | 35.0 (19.0, 48.0) | 31.0 (17.0, 66.0) | 0.948 |
|  | Number of symptoms, median (range) | 3.0 (3.0, 3.0) | 5.0 (2.0, 10.0) | 0.127 |
|  | Symptom interval in days, median (range) | 12.0 (4.0, 15.0) | 4.0 (2.0, 11.0) | 0.077 |
|  | German is not first language | 3 (100.0%) | 16 (27.1%) | 0.026 |
|  | No. of mistakes during sampling procedure, median (range) | 2.0 (1.0, 3.0) | 0.0 (0.0, 3.0) | 0.012 |
| GW | N | 7 | 55 |  |
|  | Ct value of professionally collected OP/NP, median (IQR) | 26.6 (23.4, 28.1) | 20.1(17.1, 22.3) | < 0.001 |
|  | Female | 4 (57.1%) | 27 (49.1%) | 1.000 |
|  | Age in years, median (range) | 34.0 (19.0, 48.0) | 31.0 (17.0, 66.0) | 0.798 |
|  | Number of symptoms, median (range) | 6.0 (3.0, 9.0) | 5.0 (2.0, 10.0) | 0.794 |
|  | Symptom interval in days, median (range) | 4.0 (3.0, 15.0) | 5.0 (2.0, 11.0) | 0.804 |
|  | German is not first language | 4 (57.1%) | 15(27.3%) | 0.187 |
|  | No. of mistakes during sampling procedure, median (range) | 0.0 (0.0, 2.0) | 0.0 (0.0, 2.0) | 0.426 |

MS – Multis-swab (cheek, tongue, nares), SN – Saliva-nasal (saliva, nasal vestibule), GW – gargle water. A patient sample was considered positive when either the e-gene or the ORF1ab-gene or both were detected.

In a binomial logistic regression model fit on our data, for every day of symptom duration, the odds for a positive test decreased by 40% (OR, 0.6; 95%CI, 0.4-0.9; *P*=0.01, for both MS and SN samples). Following the model, sensitivity (as compared to oro-nasopharyngeal swabs) dropped below 90% for symptom duration longer than eight days. In addition, when only assessing patients with a symptom duration of less than 8 days in our study population, test sensitivity for SN samples was 98.2% (95%CI, 90.4, 100.0), for MS 96.4% (95%CI, 87.7, 99.6).

## Discussion

Our findings indicate that self-collected samples provide only slightly reduced sensitivity in the detection of SARS-CoV-2 by RT-PCR as compared to professional-collected oro-nasopharyngeal samples. This is particularly true for sample collection using swabs (MS, SN), for sampling in the first week of symptom onset and when operational errors are minimized using comprehensive instructions.

Acceptable sensitivity when using self-collected samples including gargling techniques has previously been reported(Fernandez-Gonzalez et al., 2021, Lee et al., 2021, Lindner et al., 2020, McCulloch et al., 2020, Migueres et al., 2020, Tu Yuan-Po et al., 2020, Tu Y.P. et al., 2020, Wehrhahn et al., 2020, Wyllie et al., 2020). Public health agencies like the US Centers for Disease Control and Prevention, the Infectious Diseases Society of America, and the German Robert Koch Institute consider self-collecting techniques for symptomatic patients as potential alternatives under certain circumstances, However, they emphasize the scarcity of available data and the potential of erroneous conduct and results(Centers for Disease Control and Prevention, 2021, Kojima et al., 2020, Robert Koch Institute, 2021).

The critical temporal roles of viral shedding and sample collection was recognized early during the pandemic(Woelfel et al., 2020, Zou et al., 2020). Our data confirm a decrease of sensitivity when upper-respiratory tract samples are collected during the second week of disease(Wyllie et al., 2020). The present data suggest that until day eight of symptom onset, self-collected samples may be similarily reliable as professional collected oro-nasopharyngeal samples with sensitivities of >98% (MS, SN), and with some reservation also for GW (sensitivity >90%).

The gargling procedure performed below expactations. It is likely that the 10 ml water used for gargling instead of 1 ml of transport medium for all other samples diluted viral material in the GW sample. However, it is recommended by others(Goldfarb et al., 2021, Kojima et al., 2020) and officially used in Austria.

Diagnostic tools in the hands of untrained people require education and comprehensive instructions. Indeed, the false negative results of the patient-collected procedures were associated with procedural errors and reduced German language competence (written instructions in German). This shows an even higher potential for sample self-collection when pictorial illustrations are offered, and in different languages.

Supervision and support of self-collection procedures may be provided directly by personnel through a window or *via* video consultation. Such would not require personal protective equipment and still reduce transmission risks at testing sites.

With respect to the slightly reduced sensitivity of self-collected samples (MS and SN) in this study, it needs to be taken into account that oro-nasopharyngeal swabbing was performed by very experienced medical professionals. In a scenario of massive up-scale of testing by public health systems this may not be the case, potentially shrinking the sensitivity differences between professional and self-collection of samples. This has particular significance for a central component of pandemic response globally, i.e., repetitive testing of groups, e.g., school attendees or employees as claimed also by the WHO(World Health Organization, 2021). For that, testing including sample-collection needs to be simplified, non-invasive to prevent refusal, efficient and safe. In this regard, we provide further evidence that (home) sample self-collection may help to reduce test restraints, transmission risks as well as human and material resources. This data refers only to self-collecting for RT-PCR and not to self-collecting for rapid-tests. However the results may support the reliability of self-testing based on rapid tests as public-health tool which is currently becomming the most frequent test-method in the reality.

A limitation of our study is that only symptomatic patients were included. While there is no reason to believe that self-collection does not work in asymptomatic individuals, findings with respect to, e.g., symptom duration are not transferrable. Our study among SARS-CoV-2 positive patients could not produce specificity data. However, since specificity is predominantely determined by the diagnostic assay applied, we assume that this parameter is comparable for the sampling method. Moreover, potentially decreased specificity could still be addressed by confirming positivity *via* professional sample collections.

## Conclusion

Self-collected samples based on oral and nasal swabs for the detection of SARS-CoV-2 by RT-PCR offer very similar sensitivity compared to professional-collected oronasopharyngeal samples, if symptom duration does not exceed eight days and operational errors are minimized using comprehensive instructions. Self-sampling could thus optimize and complement containment and mitigation strategies during pandemic conditions, save human and material resources, and reduce transmission risks during sample collection and attending test sites.

## Data Availability

All raw data and analysis code are available upon a request to the corresponding author.

## Acknowledgements

Mia Wintel, Julia Macos

## Author contributions to be completed

MG, AKL, UP, FPM, FH, designed the study and developed standard operating procedures. MG, UP, SB, NK, CH, FK, ON implemented the study design, enrolled patients. MG led the writing of the manuscript. EK, JM, AN were responsible for PCR testing and contributed to the interpretation of the data. FPM, MG, HR and JS coordinated and supervised the outpatient-testing center. CR, SB, FK, UP, NK and MG enrolled patients. WvL led the data analysis. All authors have reviewed the manuscript.

## Data availability

All raw data and analysis code are available upon a request to the corresponding author.

## References

Centers for Disease Control and Prevention. Interim Guidelines for Collecting and Handling of Clinical Specimens for COVID-19 Testing; 2021. Available from: https://www.cdc.gov/coronavirus/2019-nCoV/lab/guidelines-clinical-specimens.html#handling-specimens-safely. [Accessed March 31, 2021.

Corman VM, Landt O, Kaiser M, Molenkamp R, Meijer A, Chu DK, et al. Detection of 2019 novel coronavirus (2019-nCoV) by real-time RT-PCR. Euro Surveill 2020;25(3).

Fernandez-Gonzalez M, Agullo V, de la Rica A, Infante A, Carvajal M, Garcia JA, et al. Performance of saliva specimens for the molecular detection of SARS-CoV-2 in the community setting: does sample collection method matter? Journal of clinical microbiology 2021.

Goldfarb DM, Tilley P, Al-Rawahi GN, Srigley JA, Ford G, Pedersen H, et al. Self-collected Saline Gargle Samples as an Alternative to Healthcare Worker Collected Nasopharyngeal Swabs for COVID-19 Diagnosis in Outpatients. Journal of clinical microbiology 2021.

Kirchner S, Kraemer KM, Schulze M, Pauly D, Jacob D, Gessler F, et al. Pentaplexed Quantitative Real-Time PCR Assay for the Simultaneous Detection and Quantification of Botulinum Neurotoxin-Producing Clostridia in Food and Clinical Samples. Applied and Environmental Microbiology 2010;76(13):4387–95.

Kojima N, Turner F, Slepnev V, Bacelar A, Deming L, Kodeboyina S, et al. Self-Collected Oral Fluid and Nasal Swab Specimens Demonstrate Comparable Sensitivity to Clinician-Collected Nasopharyngeal Swab Specimens for the Detection of SARS-CoV-2. Clinical infectious diseases : an official publication of the Infectious Diseases Society of America 2020.

Lee RA, Herigon JC, Benedetti A, Pollock NR, Denkinger CM. Performance of Saliva, Oropharyngeal Swabs, and Nasal Swabs for SARS-CoV-2 Molecular Detection: A Systematic Review and Meta-analysis. Journal of clinical microbiology 2021.

Lindner AK, Nikolai O, Kausch F, Wintel M, Hommes F, Gertler M, et al. Head-to-head comparison of SARS-CoV-2 antigen-detecting rapid test with self-collected anterior nasal swab versus professional-collected nasopharyngeal swab. The European respiratory journal 2020.

Lu R, Zhao X, Li J, Niu P, Yang B, Wu H, et al. Genomic characterisation and epidemiology of 2019 novel coronavirus: implications for virus origins and receptor binding. Lancet 2020;395(10224):565–74.

Maechler F, Gertler M, Hermes J, van Loon W, Schwab F, Piening B, et al. Epidemiological and clinical characteristics of SARS-CoV-2 infections at a testing site in Berlin, Germany, March and April 2020-a cross-sectional study. Clin Microbiol Infect 2020;26(12):1685.e7-.e12.

Marty FM, Chen K, Verrill KA. How to Obtain a Nasopharyngeal Swab Specimen. New England Journal of Medicine 2020;382(22).

McCulloch DJ, Kim AE, Wilcox NC, Logue JK, Greninger AL, Englund JA, et al. Comparison of Unsupervised Home Self-collected Midnasal Swabs With Clinician-Collected Nasopharyngeal Swabs for Detection of SARS-CoV-2 Infection. Jama Network Open 2020;3(7).

Michel D, Danzer KM, Gross R, Conzelmann C, Muller JA, Freischmidt A, et al. Rapid, convenient and efficient kit-independent detection of SARS-CoV-2 RNA. J Virol Methods 2020;286.

Migueres M, Mengelle C, Dimeglio C, Didier A, Alvarez M, Delotel P, et al. Saliva sampling for diagnosing SARS-CoV-2 infections in symptomatic patients and asymptomatic carriers. Journal of Clinical Virology 2020;130.

Pan Y, Zhang D, Yang P, Poon LLM, Wang Q. Viral load of SARS-CoV-2 in clinical samples. Lancet Infect Dis 2020;20(4):411–2.

Rai P, Kumar BK, Deekshit VK, Karunasagar I, Karunasagar I. Detection technologies and recent developments in the diagnosis of COVID-19 infection. Applied Microbiology and Biotechnology 2021;105(2):441–55.

Robert Koch Institute. Hinweise zur Testung von Patienten auf Infektion mit dem neuartigen Coronavirus SARS-CoV-2; 2021. Available from: https://www.rki.de/DE/Content/InfAZ/N/Neuartiges_Coronavirus/Vorl_Testung_nCoV.html;jsessionid=CD421678C21E595557DF2DB07E1E590A.internet072?nn=13490888#doc13490982bodyText1. [Accessed March 31, 2021 2021].

Tu Y-P, Jennings R, Berke EM. Swabs Collected by Patients or Health Care Workers for SARS-CoV-2 Testing. New England Journal of Medicine 2020;383(5):494–6.

Tu YP, Jennings R, Hart B, Cangelosi GA, Wood RC, Wehber K, et al. Patient-collected tongue, nasal, and mid-turbinate swabs for SARS-CoV-2 yield equivalent sensitivity to health care worker collected nasopharyngeal swabs. 2020.

Wehrhahn MC, Robson J, Brown S, Bursle E, Byrne S, New D, et al. Self-collection: An appropriate alternative during the SARS-CoV-2 pandemic. Journal of clinical virology : the official publication of the Pan American Society for Clinical Virology 2020;128:104417-.

Woelfel R, Corman VM, Guggemos W, Seilmaier M, Zange S, Mueller MA, et al. Virological assessment of hospitalized patients with COVID-2019. Nature 2020;581(7809):465-+.

World Health Organization. COVID-19 strategic preparedness and response plan: 1 February 2021 to 31 January 2022. Geneva: World Health Organization; 2021.

Wyllie AL, Fournier J, Casanovas-Massana A, Campbell M, Tokuyama M, Vijayakumar P, et al. Saliva or Nasopharyngeal Swab Specimens for Detection of SARS-CoV-2. New England Journal of Medicine 2020;383(13):1283–6.

Zhu N, Wong PK. Advances in Viral Diagnostic Technologies for Combating COVID-19 and Future Pandemics. Slas Technology 2020;25(6):513–21.

Zhu N, Zhang D, Wang W, Li X, Yang B, Song J, et al. A Novel Coronavirus from Patients with Pneumonia in China, 2019. New England Journal of Medicine 2020;382(8):727–33.

Zou L, Ruan F, Huang M, Liang L, Huang H, Hong Z, et al. SARS-CoV-2 Viral Load in Upper Respiratory Specimens of Infected Patients. New England Journal of Medicine 2020;382(12):1177–9.

